# Bivalent COVID-19 vaccine antibody responses to Omicron variants suggest that responses to divergent variants would be improved with matched vaccine antigens

**DOI:** 10.1101/2023.02.22.23286320

**Authors:** Wei Wang, Emilie Goguet, Stephanie Paz Padilla, Russell Vassell, Simon Pollett, Edward Mitre, Carol D. Weiss

**Affiliations:** Center for Biologics Evaluation and Research, US Food and Drug Administration, Silver Spring, MD, USA; Department of Microbiology and Immunology, Uniformed Services University of the Health Sciences, Bethesda, MD, USA; Infectious Diseases Clinical Research Program, Department of Preventive Medicine and Biostatistics, Uniformed Services University of the Health Sciences, Bethesda, MD, USA; Henry M. Jackson Foundation for the Advancement of Military Medicine, Inc., Bethesda, Maryland, United States of America

## Abstract

We compared neutralizing antibody responses to BA.4/5, BQ.1.1, XBB, and XBB.1.5 Omicron SARS-CoV-2 variants after a bivalent or ancestral COVID-19 mRNA booster vaccine or post-vaccination infection. We found that the bivalent booster elicited moderately high antibody titers against BA.4/5 that were approximately two-fold higher against all Omicron variants than titers elicited by the monovalent booster. The bivalent booster elicited low but similar titers against both XBB and XBB.1.5 variants. These findings inform risk assessments for future COVID-19 vaccine recommendations and suggest that updated COVID-19 vaccines containing matched vaccine antigens to circulating divergent variants may be needed.

## Introduction

The evolution of severe acute respiratory syndrome coronavirus 2 (SARS-CoV-2) and dynamic population immunity from combinations of vaccines and infections present challenges for vaccination strategies. Since the Omicron BA.5 subvariant became dominant in mid-2022, additional immune-evasive Omicron variants emerged, including BA.5 lineage BQ.1 and BQ.1.1 variants and BA.2 lineage recombinant variants XBB, XBB.1 and XBB.1.5 (Fig. 1A). Bivalent mRNA COVID-19 vaccines encoding both ancestral (D614G) and Omicron BA.4/5 spike proteins have been used to boost immunity since September 2022. However, there are concerns that prior monovalent vaccinations encoding the ancestral variant may hinder antibody responses to new variants through immune imprinting [1, 2]. Recommendations about the timing of COVID-19 vaccine boosters and variant composition of updated COVID-19 vaccines are based on risk-assessments that consider pre-existing immunity. Understanding how bivalent boosting compares to monovalent ancestral boosting or a post-vaccine infection (PVI) in eliciting neutralizing antibodies to recent circulating Omicron variants, including XBB and XBB.1.5, informs risk-assessments for decisions about use of vaccine boosters or vaccine composition updates [3].

**Figure 1.**
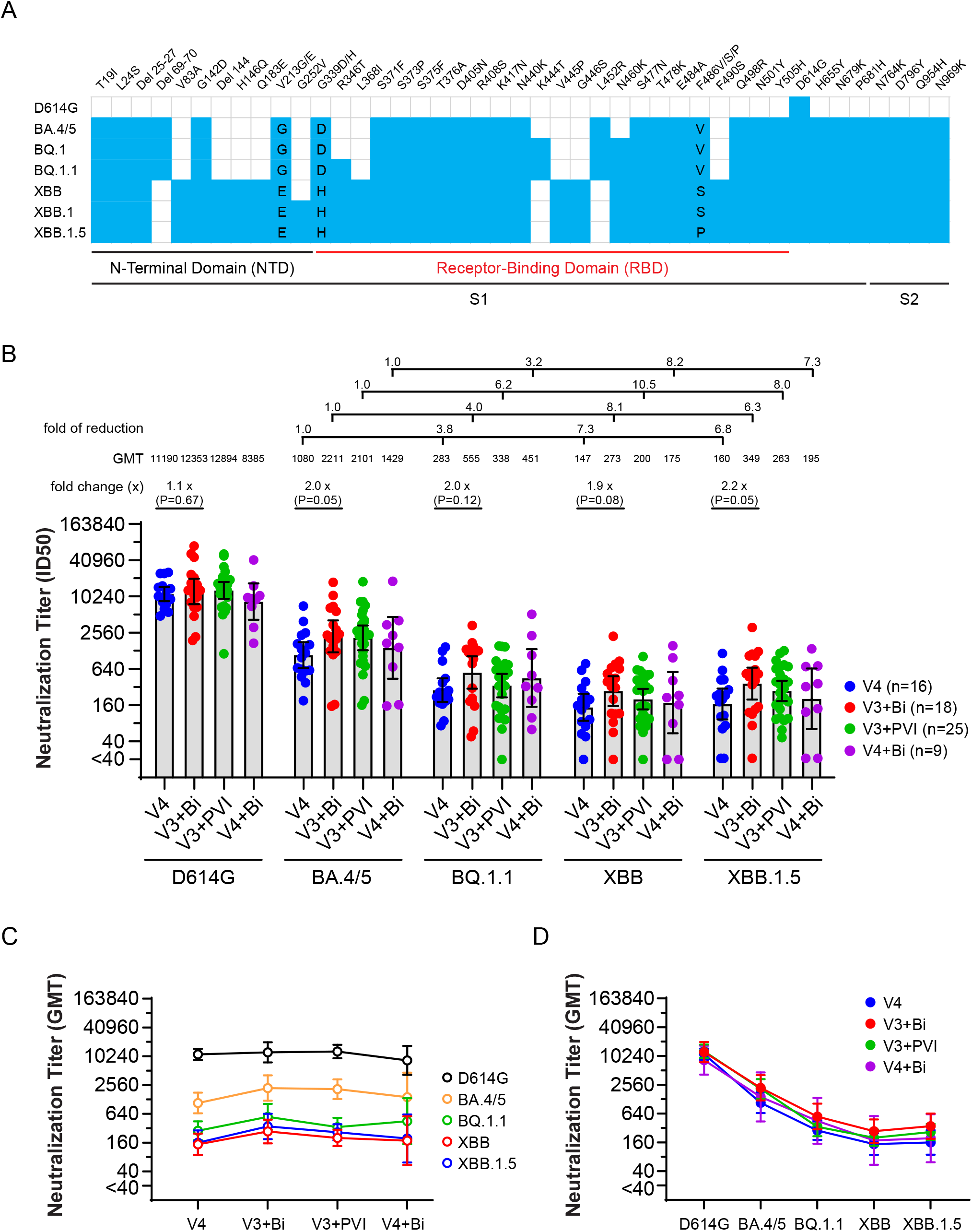
Neutralization of Omicron Subvariants by Post-vaccination and Post-vaccination Infection Serum Samples. **A.** Amino acid mutations and deletions (Del) in spike proteins of ancestral (D614G) and recently emerged Omicron sub-variants are indicated in reference to the USA-WA1/2020. Blue boxes indicate an amino acid substitution relative to WA1/2020. Amino acid substitutions, indicated by their single letter abbreviation, are listed in the blue box for variants that have different substitutions in those positions. N-terminal domain (NTD) and receptor binding domain (RBD) in S1 are marked. BA.4 and BA.5 have the identical spike sequence, thus their spikes were marked as BA.4/5. **B.** Neutralizing antibody titers against the indicated variants in human serum samples after different exposures by mRNA COVID-19 vaccines and post-vaccination infections were measured in lentiviral-based pseudovirus neutralization assays. Geometric mean titers (GMTs) assessed as the reciprocal dilution of serum that neutralizes 50% of the input pseudovirus (ID_50_) against different variant pseudoviruses were compared. Serum samples in which the ID_50_ fell below the limit of detection at 1:40 dilution were assigned an ID_50_ value of 20. Dots indicate results from individual participants, and bars indicate GMT with 95% confidence interval. P values of less than 0.05 were considered statistically significant. All neutralization titers were log2 transformed for analyses. **C.** The neutralizing antibody titers (GMT) in V4, V3+Bi, V3+PVI and V4+Bi groups are illustrated with titers against D614G, BA.4/5, BQ.1.1, XBB and XBB.1.5. **D.** The neutralizing antibody titers (GMT) against D614G, BA.4/5, BQ.1.1, XBB and XBB.1.5 are compared in V4, V3+Bi, V3+PVI and V4+Bi groups.

## Methods

### Ethics and study cohort

Serum samples used in this study were obtained from participants in the Prospective Assessment of SARS-CoV-2 Seroconversion study (PASS study) that was approved by the Uniformed Services University of the Health Sciences Institutional Review Board (Federalwide Assurance no. 00001628, US Department of Defense Assurance no. P60001) in compliance with all applicable federal regulations governing the protection of human participants. Written informed consent was obtained from all study participants. We measured serum neutralizing antibody titers against lentiviral pseudoviruses with D614G, BA.4/5, BQ.1.1, XBB and XBB.1.5 spike proteins from persons with different exposure histories in a well-characterized, prospective cohort (see Supplementary Figure 1 and Supplementary Table 1). Serum from individuals who received four ancestral mRNA vaccines (V4, n=16) was collected 7-73 (median 33) days after last vaccination. Serum from individuals who received three ancestral and one bivalent mRNA vaccine (V3+Bi, n=18) was collected 8-65 (median 29) days after last vaccination. Serum from individuals who received three ancestral mRNA vaccines before a PVI during the BA.1 wave (V3+PVI, n=25) were collected 7-99 (median 56) days after infection. We also collected serum from persons who received four doses of ancestral mRNA vaccine before one bivalent mRNA vaccine (V4+Bi, n=9) at 20-55 (median 35) days after last vaccination.

### Pseudovirus production and neutralization assay

HIV-based lentiviral pseudoviruses with desired SARS-CoV-2 spike proteins (D614G, BA.4/5, BQ.1.1, XBB and XBB.1.5) were generated as previously described [4]. Pseudoviruses were produced in 293T cells by co-transfection of 5 mg of pCMVΔR8.2, 5 mg of pHR’CMVLuc and 0.5 mg of pVRC8400 encoding a codon-optimized spike gene. Pseudovirus supernatants were collected approximately 48 h post transfection, filtered through a 0.45 mm low protein binding filter, and stored at -80°C. Pseudovirus neutralization assays were performed using 293T-ACE2-TMPRSS2 cells in 96-well plates [4]. Pseudoviruses with titers of approximately 10^6^ relative luminescence units per milliliter (RLU/mL) of luciferase activity were incubated with serially diluted sera for two hours at 37°C prior to inoculation onto the plates that were pre-seeded one day earlier with 3.0 × 10^4^ cells/well. Pseudovirus infectivity was determined 48 h post inoculation for luciferase activity by luciferase assay reagent (Promega) according to the manufacturer’s instructions. The inverse of the sera dilutions causing a 50% reduction of RLU compared to control was reported as the neutralization titer (ID_50_). Titers were calculated using a nonlinear regression curve fit (GraphPad Prism Software Inc., La Jolla, CA, USA). The mean titer from at least two independent experiments each with intra-assay duplicates was reported as the final titer.

### Statistics analysis

Mann Whitney test for two group comparisons, Tukey’s multiple comparisons test for multiple groups and geometric mean titers (GMT) with 95% confidence intervals were performed using GraphPad Prism software. The *p* values of less than 0.05 were considered statistically significant. All neutralization titers were log_2_ transformed for analyses.

## Results

The primary outcome assessed in this study was neutralizing titers between V4 and V3+Bi groups. Neutralizing antibody titers in individuals who received a 4^th^ vaccine dose with bivalent vaccine (V3+Bi) were approximately two-fold greater against BA.4/5, BQ.1.1, XBB and XBB.1.5 (P values 0.05, 0.12, 0.08 and 0.05, respectively) than titers in individuals who received four doses of ancestral mRNA vaccine (V4) (Fig. 1B). We also compared neutralizing antibody titers against individual variants between each of the antigen exposure groups (V4 versus V3+Bi versus V3+PVI versus V4+Bi). In this multiple comparison analysis, there were no significant differences for D614G, BA.4/5, BQ.1.1, XBB and XBB.1.5. Within each V4, V3+Bi, V3+PVI, V4+Bi group, only D614G versus BA.4/5, BQ.1.1, XBB and XBB.1.5 have significant differences (all *P*<0.01). Overall, GMTs against D614G were similar among the groups (Fig. 1B and 1C). Notably, GMTs against BQ.1.1, XBB.1 and XBB.1.5 were low at levels 3 to 11-fold lower than GMTs against BA.4/5. GMTs against XBB and XBB.1.5 were similar within each exposure group (Fig.1B and 1C). GMTs in the V3+Bi and V3+PVI groups were similar, while the V4+Bi group had modestly lower GMTs (Fig. 1B and 1D).

## Discussion

Although concerns about immune imprinting from prior ancestral vaccine exposures remain [5], we found that a booster dose of bivalent vaccine (V3+Bi) elicited modestly higher titers against most recent Omicron variants, including XBB and XBB.1.5 variants, than four doses of ancestral vaccine (V4). Within any antigen-exposure group, titers against XBB and XBB.1.5 were similar, though considerably lower than against BA.4/5, indicating significant antigenic divergence. Our neutralizing antibody results align with clinical data showing increased vaccine effectiveness of bivalent boosting against BA.5– and XBB/XBB.1.5–related infections compared to monovalent boosting [6, 7]. As most PVIs occurred when Omicron variants predominated, GMTs from V3+Bi, V3+PVI, and V4+Bi groups (Fig. 1B and 1D) suggest that boosting by an infection or vaccine that more closely matches circulating variants may be better than boosting by the ancestral variant. The modest increase in titers against BA.4/5 after the bivalent booster or Omicron infection may represent a priming response to novel epitopes in the BA.4/5 spike that are not present in the ancestral spike. The current bivalent booster vaccine contains mRNAs encoding a half-dose each of the ancestral and BA.4/5 spikes. Whether a first monovalent booster with a full dose of an Omicron variant, a second bivalent booster, or a subsequent Omicron exposure would improve titers to Omicron variants remains to be determined.

Nonetheless, because the bivalent vaccine and PVI groups elicited strong boosting to the prior D614G variant, evaluation of booster vaccines including only new variants appears warranted. Further, to improve protection against variants with high antigenic divergence from the vaccine antigens, such as XBB and XBB.1.5, updated COVID-19 vaccines containing matched antigens to divergent variants may be needed. Study limitations include small sample numbers, a relatively healthy cohort, modestly different median ages among the groups, potential asymptomatic infections, and non-contemporaneous sample collections. Study strengths include well-characterized vaccinees followed prospectively and a broad number of Omicron variants tested.

## Supporting information

XBB-Wangetal-fullsupplement-medrxiv

## Data Availability

Data are available in Supplementary Tables 1 and 2.

## Acknowledgements

The authors gratefully acknowledge all research volunteers for their time and participation and all PASS study team members for their contributions to the PASS study.

## Supplementary Data

Supplementary Figure 1, Supplementary Table 1, Supplementary Table 2

### Funding/Support

The protocol was executed by the Infectious Disease Clinical Research Program (IDCRP), a Department of Defense (DoD) program executed by the Uniformed Services University of the Health Sciences (USUHS) through a cooperative agreement by the Henry M. Jackson Foundation for the Advancement of Military Medicine, Inc. (HJF). This work was supported in whole, or in part, with federal funds from the US Food and Drug Administration Medical Countermeasures Initiative grant # OCET 2022-1750, and the Defense Health Program (HU00012020067, HU00012020094) and the Immunization Healthcare Branch (HU00012120104) of the Defense Health Agency, United States Department of Defense, and the National Institute of Allergy and Infectious Disease (HU00011920111), under Inter-Agency Agreement Y1-AI-5072.

### Data Sharing Statement

Data are available in Supplementary Tables 1 and 2. Questions or additional requests should be directed to the communicating author.

### Author contributions

#### Concept and design

Wang, Pollett, Mitre, Weiss *Acquisition, analysis, or interpretation of data:* All authors.

#### Drafting of the manuscript

Wang, Weiss.

#### Critical revision of the manuscript for important intellectual content

All authors.

#### Statistical analysis

Wang

#### Obtained funding

Mitre, Weiss

#### Administrative, technical, or material support

Paz Padilla, Vassell

#### Supervision

Pollett, Mitre, Weiss

### Disclaimer

The contents of this publication are the sole responsibility of the author (s) and do not necessarily reflect the views, opinions, or policies of the Food and Drug Administration; Uniformed Services University of the Health Sciences (USUHS); the Department of Defense (DoD); the Departments of the Army, Navy, or Air Force; the Defense Health Agency; WRNMMC; and the Henry M. Jackson Foundation for the Advancement of Military Medicine Inc. Mention of trade names, commercial products, or organizations does not imply endorsement by the U.S. Government. The investigators have adhered to the policies for protection of human subjects as prescribed in 45 CFR 46.

Paz Padilla, Vassell, Wang, Mitre, and Weiss are employees of the US Government. This work was prepared as part of their official duties. Title 17 USC §105 provides that copyright protection under this title is not available for any work of the US Government. Title 17 USC §101 defines a US Government work as a work prepared by a military service member or employee of the US Government as part of that person’s official duties.

Dr Pollett is employed by the Henry M. Jackson Foundation for the Advancement of Military Medicine Inc.

