## Supplementary material for "Bivalent COVID-19 vaccine antibody responses to Omicron variants suggest that responses to divergent variants would be improved with matched vaccine antigens": XBB-Wangetal-fullsupplement-medrxiv

#### Supplementary Data

Supplementary Figure 1, Supplementary Table 1, Supplementary Table 2

### Bivalent COVID-19 vaccine antibody responses to Omicron variants suggest that responses to divergent variants would be improved with matched vaccine antigens

Wei Wang[1], Emilie Goguet[2], Stephanie Paz Padilla[1], Russell Vassell[1], Simon Pollett[3,4], Edward Mitre[2*], Carol D. Weiss[1*]

**Affiliations**

[1]Center for Biologics Evaluation and Research, US Food and Drug Administration, Silver Spring, MD, USA

[2]Department of Microbiology and Immunology, Uniformed Services University of the Health Sciences, Bethesda, MD, USA

[3]Infectious Diseases Clinical Research Program, Department of Preventive Medicine and Biostatistics, Uniformed Services University of the Health Sciences, Bethesda, MD, USA

[4]Henry M. Jackson Foundation for the Advancement of Military Medicine, Inc., Bethesda, Maryland, United States of America

Supplementary Figure 1

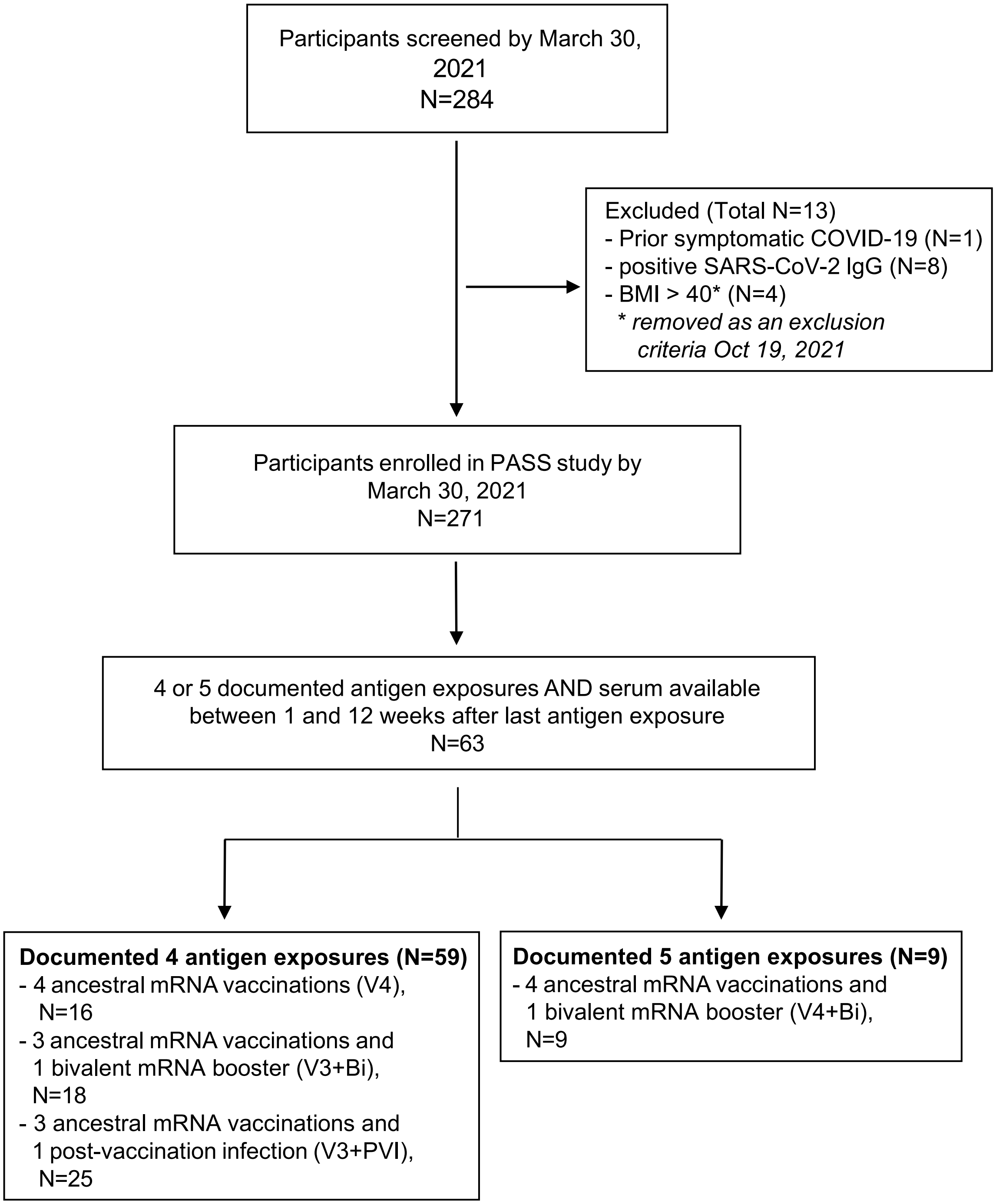

**Supplementary Figure 1. Strobe chart of the study cohort.** Details of the Prospective Assessment of SARS-CoV-2 Seroconversion (PASS) study protocol, including details of the inclusion/exclusion criteria, have been published (Jackson-Thompson BM, Goguet E, Laing ED, et al. Prospective Assessment of SARS-CoV-2 Seroconversion (PASS) study: an observational cohort study of SARS-CoV-2 infection and vaccination in healthcare workers. *BMC Infect Dis*. Jun 9 2021;21(1):544. doi:10.1186/s12879-021-06233-1). The PASS (Protocol IDCRP-126) studies were approved by the Uniformed Services University of the Health Sciences Institutional Review Board in compliance with all applicable federal regulations governing the protection of human participants. All study participants provided informed consent. Inclusion criteria included being generally healthy, ≥ 18 years old, and employed at the Walter Reed National Military Medical Center. Exclusion criteria included history of COVID-19, IgG seropositivity for SARS-CoV-2, and being severely immunocompromised at time of screening. The study was initiated in August 2020, with rolling enrollment through February of 2021. Serum samples for antibody testing were collected monthly through August of 2021, then quarterly through August of 2022, and then semiannually after that. Participants are asked to test for SARS-CoV-2 infection any time they have symptoms. Details of SARS-CoV-2 infections and vaccinations are collected at every study visit. A total of 271 individuals were enrolled, with 148 still being actively followed as of January 2023. The subset of participants included in this study were those who had four or five documented antigen exposures (sum of vaccine doses and SARS-CoV-2 infections) and had serum available between one and 12 weeks after the most recent antigen exposure. A total of 63 participants were included in this study, and a total of 68 serum samples were analyzed as five of the participants had serum available after both four and five antigen exposures. N: number of participants.

**Supplementary Table 1**. **Characteristics of clinical cohort.**

| Characteristics | Overall population (N=63) | V4 (N=16) | V3+Bi (N=18) | V3+PVI (N=25) | V4+Bi (N=9) |
|---|---|---|---|---|---|
| **Sex** | | | | | |
| Female | 42 (66.7%) | 10 (62.5%) | 10 (55.6%) | 18 (72.0%) | 7 (77.8%) |
| Male | 21 (33.3%) | 6 (37.5%) | 8 (44.4%) | 7 (28.0%) | 2 (22.2%) |
| **Age** | | | | | |
| 18-44 | 29 (46.0%) | 2 (12.5%) | 14 (77.8%) | 12 (48.0%) | 1 (11.1%) |
| 45-64 | 32 (50.8%) | 13 (81.3%) | 4 (22.2%) | 13 (52.0%) | 7 (77.8%) |
| 65+ | 2 (3.2%) | 1 (6.2%) | 0 (0%) | 0 (0%) | 1 (11.1%) |
| Median age (IQR) | 46 (16) | 55.5 (7) | 39.5 (7.75) | 45 (15) | 54 (6) |
| **Charlson Co-morbidity Index** | | | | | |
| 0 | 53 (84.1%) | 13 (81.3%) | 16 (88.9%) | 21 (84.0%) | 6 (66.7%) |
| 1 | 7 (11.1%) | 1 (6.2%) | 2 (11.1%) | 3 (12.0%) | 1 (11.1%) |
| 2 | 2 (3.2%) | 1 (6.2%) | 0 (0%) | 1 (4.0%) | 1 (11.1%) |
| 3 | 1 (1.6%) | 1 (6.2%) | 0 (0%) | 0 (0%) | 1 (11.1%) |
| **Length of time (days) between serum collection and most recent antigen exposure** | | | | | |
| Median days (IQR) | 35 (29) | 33 (30.5) | 29 (7) | 56 (38) | 35 (14) |

Sixty-three participants were included in this study and a total of 68 serum samples were collected and distributed into four antigen exposure groups: V3+Bi (3 ancestral mRNA and 1 bivalent mRNA vaccines), V3+PVI (3 ancestral mRNA vaccines followed by a post-vaccination infection during BA.1 wave), V4 (4 ancestral mRNA vaccines), and V4+Bi (4 ancestral mRNA and 1 bivalent mRNA vaccines. Serum was collected at two different antigen exposure timepoints for five participants who contributed to the V4 and V4+Bi groups. IQR, Interquartile range.

#### Supplementary Table 2. Neutralization titers

| ID | Participant status at time of collection | Time between last vax and collection (days) | Time between infection and collection (days) | D614G | BQ.1.1 | BA.4/5 | XBB | XBB.1.5 |
|---|---|---|---|---|---|---|---|---|
| 01 | V4 | 7 | | 14471 | 2314 | 375 | 133 | 174 |
| 02 | V4 | 51 | | 8322 | 589 | 186 | 199 | 217 |
| 03 | V4 | 10 | | 13786 | 1613 | 279 | 157 | 233 |
| 04 | V4 | 27 | | 24577 | 7071 | 1477 | 606 | 1100 |
| 05 | V4 | 23 | | 12634 | 376 | 87 | 92 | 98 |
| 06 | V4 | 21 | | 15268 | 1365 | 218 | 164 | 191 |
| 07 | V4 | 40 | | 4841 | 189 | 73 | 53 | <40 |
| 08 | V4 | 56 | | 9989 | 1663 | 227 | 48 | 62 |
| 09 | V4 | 14 | | 9930 | 987 | 442 | 230 | 274 |
| 10 | V4 | 52 | | 9138 | 1177 | 198 | 85 | 103 |
| 11 | V4 | 32 | | 10273 | 2536 | 1268 | 781 | 402 |
| 12 | V4 | 25 | | 5593 | 664 | 258 | 44 | <40 |
| 13 | V4 | 67 | | 6313 | 604 | 166 | 196 | 222 |
| 14 | V4 | 57 | | 7661 | 470 | 174 | 71 | 70 |
| 15 | V4 | 34 | | 25657 | 3916 | 867 | 392 | 577 |
| 16 | V4 | 73 | | 24528 | 632 | 262 | 304 | 403 |
| 17 | V3+Bi | 35 | | 9927 | 2207 | 1356 | 530 | 451 |
| 18 | V3+Bi | 18 | | 2180 | 153 | 59 | <40 | <40 |
| 19 | V3+Bi | 28 | | 23643 | 3447 | 1123 | 440 | 1181 |
| 20 | V3+Bi | 28 | | 6752 | 2060 | 209 | 116 | 115 |

| | | | | | | | | |
|---|---|---|---|---|---|---|---|---|
| 21 | V3+Bi | 27 | | 12846 | 2312 | 1345 | 445 | 592 |
| 22 | V3+Bi | 29 | | 51977 | 6948 | 1735 | 311 | 542 |
| 23 | V3+Bi | 8 | | 10807 | 1267 | 154 | 115 | 144 |
| 24 | V3+Bi | 27 | | 20919 | 2412 | 1277 | 639 | 502 |
| 25 | V3+Bi | 42 | | 70398 | 17467 | 3364 | 2226 | 2980 |
| 26 | V3+Bi | 33 | | 45749 | 5806 | 939 | 392 | 507 |
| 27 | V3+Bi | 39 | | 15990 | 6573 | 1194 | 669 | 1010 |
| 28 | V3+Bi | 35 | | 8213 | 1985 | 308 | 114 | 108 |
| 29 | V3+Bi | 65 | | 6775 | 1098 | 191 | 200 | 333 |
| 30 | V3+Bi | 29 | | 16935 | 10600 | 1517 | 861 | 998 |
| 31 | V3+Bi | 28 | | 4799 | 1230 | 184 | 83 | 129 |
| 32 | V3+Bi | 33 | | 1890 | 163 | 48 | 56 | 70 |
| 33 | V3+Bi | 28 | | 15191 | 2854 | 779 | 765 | 1077 |
| 34 | V3+Bi | 54 | | 12858 | 1532 | 1253 | 324 | 391 |
| 35 | V3+PVI | 264 | 91 | 31776 | 8335 | 614 | 350 | 411 |
| 36 | V3+PVI | 123 | 7 | 6567 | 189 | <40 | <40 | 44 |
| 37 | V3+PVI | 286 | 63 | 11339 | 2361 | 469 | 294 | 373 |
| 38 | V3+PVI | 120 | 27 | 26460 | 2822 | 652 | 1026 | 712 |
| 39 | V3+PVI | 139 | 37 | 48306 | 8362 | 1484 | 618 | 1225 |
| 40 | V3+PVI | 123 | 42 | 11954 | 1238 | 334 | 152 | 201 |
| 41 | V3+PVI | 169 | 71 | 16645 | 2259 | 561 | 481 | 371 |
| 42 | V3+PVI | 267 | 40 | 21525 | 2830 | 732 | 324 | 495 |
| 43 | V3+PVI | 182 | 79 | 12641 | 1694 | 243 | 94 | 131 |
| 44 | V3+PVI | 177 | 99 | 5087 | 714 | 128 | 95 | 91 |
| 45 | V3+PVI | 328 | 93 | 15099 | 5907 | 1510 | 516 | 614 |
| 46 | V3+PVI | 167 | 80 | 16113 | 3529 | 137 | 99 | 154 |
| 47 | V3+PVI | 242 | 29 | 9934 | 511 | 148 | 71 | 89 |
| 48 | V3+PVI | 167 | 83 | 14151 | 2737 | 332 | 283 | 287 |

| | | | | | | | | |
|---|---|---|---|---|---|---|---|---|
| 49 | V3+PVI | 125 | 39 | 11190 | 881 | 97 | 71 | 85 |
| 50 | V3+PVI | 148 | 70 | 16742 | 1665 | 368 | 271 | 335 |
| 51 | V3+PVI | 160 | 70 | 16188 | 5048 | 747 | 418 | 662 |
| 52 | V3+PVI | 150 | 52 | 5833 | 796 | 64 | 84 | 94 |
| 53 | V3+PVI | 383 | 30 | 6796 | 3905 | 494 | 157 | 178 |
| 54 | V3+PVI | 169 | 22 | 1136 | 159 | 91 | 56 | 59 |
| 55 | V3+PVI | 284 | 62 | 18961 | 4144 | 776 | 382 | 847 |
| 56 | V3+PVI | 289 | 37 | 11027 | 2274 | 865 | 532 | 668 |
| 57 | V3+PVI | 245 | 23 | 7079 | 1041 | 157 | 115 | 174 |
| 58 | V3+PVI | 390 | 56 | 51643 | 17884 | 1538 | 595 | 1111 |
| 59 | V3+PVI | 138 | 75 | 19321 | 7278 | 542 | 149 | 245 |
| 60 | V4+Bi | 39 | | 1701 | 162 | 98 | <40 | <40 |
| 61 | V4+Bi | 43 | | 10494 | 1700 | 328 | 214 | 347 |
| 62 | V4+Bi | 29 | | 15316 | 2298 | 503 | 231 | 309 |
| 63 | V4+Bi | 35 | | 41391 | 18221 | 5163 | 1551 | 1320 |
| 64 | V4+Bi | 34 | | 8988 | 1455 | 309 | 162 | 117 |
| 65 | V4+Bi | 27 | | 3133 | 155 | 63 | <40 | <40 |
| 66 | V4+Bi | 55 | | 9799 | 4039 | 2279 | 975 | 683 |
| 67 | V4+Bi | 20 | | 9427 | 3448 | 1076 | 409 | 669 |
| 68 | V4+Bi | 43 | | 6959 | 687 | 193 | 79 | 134 |
